# Transition probabilities for dental caries in a school-based prevention program: A randomized clinical trial

**DOI:** 10.1101/2024.06.05.24308501

**Authors:** Ryan Richard Ruff

## Abstract

**Background:** School-based caries prevention using silver diamine fluoride (SDF) has been shown to effectively prevent and control dental caries. To better inform program design and implementation, this paper estimated transition probabilities for dental caries in a school SDF program.

**Methods:** The CariedAway project was a pragmatic, cluster-randomized trial of school-based caries prevention interventions conducted in predominately low-income minority children. For children in CariedAway receiving SDF, transition probabilities were computed between sound, carious, and arrested states for 6-year molars using multistate Markov models. Subject-level transition probabilities over one- and two-year periods were then calculated by aggregating states of all 6-year molars and first and second bicuspids.

**Results:** A total of 7418 children were enrolled in CariedAway, of which 1352 met inclusion criteria for this study. Of eligible participants, the baseline prevalence of untreated decay was 29% and the prevalence of dental sealants was 8%. The probability of transitioning between sound and carious states in 6-year molars ranged from 0.0022 to 0.0074. At the subject-level, the sound to carious transition probabilities were 0.07 and 0.12 after one and two years, respectively. Once in a fully arrested state, the probability of remaining arrested was 0.72 and 0.60 after one and two years.

**Conclusions:** The overall probabilities of teeth remaining in diseased-free or arrested states was high after receiving silver diamine fluoride, although multiple applications might be needed for consistent caries arrest.

## Introduction

Dental caries is a highly prevalent and inequitable public health crisis ^1^, negatively affecting quality of life ^2^ and educational performance ^3^. The disproportionate burden of caries often stems from a lack of access to affordable dental care ^4^. School-based caries prevention can increase access to dental services, providing cost-effective and cost-saving interventions to disadvantaged children ^5^. Known approaches to school caries prevention include dental sealants ^6^, fluoride varnish ^7^, toothbrushing and outpatient services ^8^, and atraumatic restorations ^9^, each of which has demonstrated effectiveness in reducing the risk of caries.

Silver diamine fluoride (SDF) is recommended by the American Academy of Pediatric dentistry a cost-effective approach for comprehensive caries management in children ^10^, and has published guidelines for provision by primary care providers ^11^. Available in multiple concentrations, prior studies indicate that a 38% SDF solution is the most efficacious, and arrest rates increase with semiannual application ^12^. It is also effective in the prevention of new caries ^13^. When used as part of a school-based program, SDF has been shown to be comparable to dental sealants in the prevention ^14^ and control ^15^ of dental caries, can be provided by school nurses ^16^, and can minimize the time required for children to be out of class ^17^.

The large-scale use of silver diamine fluoride for school-based caries prevention has the potential to dramatically increase the reach and effectiveness of care, and understanding anticipated patient needs can better support program planning and sustainability. Identifying the likelihood of individuals developing new caries, post-initial treatment, can help inform the scheduling of subsequent school visits and maximize the utility of limited resources. In this paper, we estimate the transition probabilities of teeth treated with SDF in a school-based caries prevention program using multistate Markov models, focusing on a traditionally underserved population consisting of predominately low-income minority 1children with both a high baseline prevalence of caries and a low prevalence of dental sealants.

## Methods

### Design and Participants

CariedAway was a cluster-randomized, longitudinal, pragmatic clinical trial that investigated the feasibility of using silver diamine fluoride in school-based caries prevention ^18^. The CariedAway study received approval from the New York University School of Medicine Institutional Review Board and is registered at clinicaltrials.gov (#NCT03442309). CariedAway was conducted from February 1 2019 to June 1 2023 in 48 primary schools located in New York City with a total student population consisting of at least 50% Black or Hispanic students and at least 80% of students receiving free or reduced lunch. Any child with parental informed consent and child assent was eligible to receive care.

### Interventions

Each study participant received either silver diamine fluoride followed by fluoride varnish (to mask the bitter aftertaste of SDF) or dental sealants, atraumatic restorations, and fluoride varnish. For the SDF group, one to two drops of SDF were dispensed into a mixing well and applied with a microapplicator to all pits and fissures of premolars and molars. SDF was also applied to all posterior asymptomatic cavitated lesions. One unit dose of fluoride varnish was then applied. For the sealant group, cavity conditioner was first applied followed by application of glass ionomer sealants using the finger-sweep technique to all pits and fissures of bicuspids and molars. Atraumatic restorations were then placed on any asymptomatic cavitated lesions, and fluoride varnish was then applied to all teeth. SDF was reapplied at every subsequent observation, and sealants were reapplied to any unsealed or partially sealed molars and bicuspids.

### Randomization and Blinding

Children were block randomized at the school level using a random number generator. As a pragmatic trial, participants were unable to be blinded.

### Examination and Diagnosis

Each participant in the CariedAway trial received a full visual-tactile examination prior to receiving the assigned interventions. Dental caries diagnosis followed the International Caries Detection and Assessment System (ICDAS). Any tooth with lesions determined to meet ICDAS score of 5 (distinct cavity with visible dentin) or 6 (extensive, more than half the surface, distinct cavity with visible dentin) was classified as decay. Arrest was classified as frank absence of tooth structure to the level of the dentin, the surface of which is black, with or without shine, that was hard and smooth when moving an explorer tip gently across the surface. Otherwise, teeth were assessed as intact/sound, sealed, or restored (e.g., silver or porcelain filling).

### Impact of COVID-19

The original protocol for CariedAway stipulated biannual examination and treatment. However, as a school-based trial, study activities were suspended from March 2020 through September 2021 due to the COVID-19 pandemic. As a result, some children enrolled in CariedAway had approximately two years before their first follow-up observation. Upon resumption of study procedures following the lifting of pandemic restrictions, the biannual schedule was resumed.

### Statistical Analysis

Using longitudinal data from the CariedAway trial, we estimated transition probabilities via multi-state Markov models in those children assigned to the SDF arm who completed at least two follow-up observations. Tooth state progression was treated as a relapsing-remitting condition consisting of discrete state transitions from sound to carious, carious to arrested, or arrested to carious (Figure 1), reflecting both primary and secondary effects of silver diamine fluoride treatment and the intermittently observed nature of the data.

**Figure 1:**
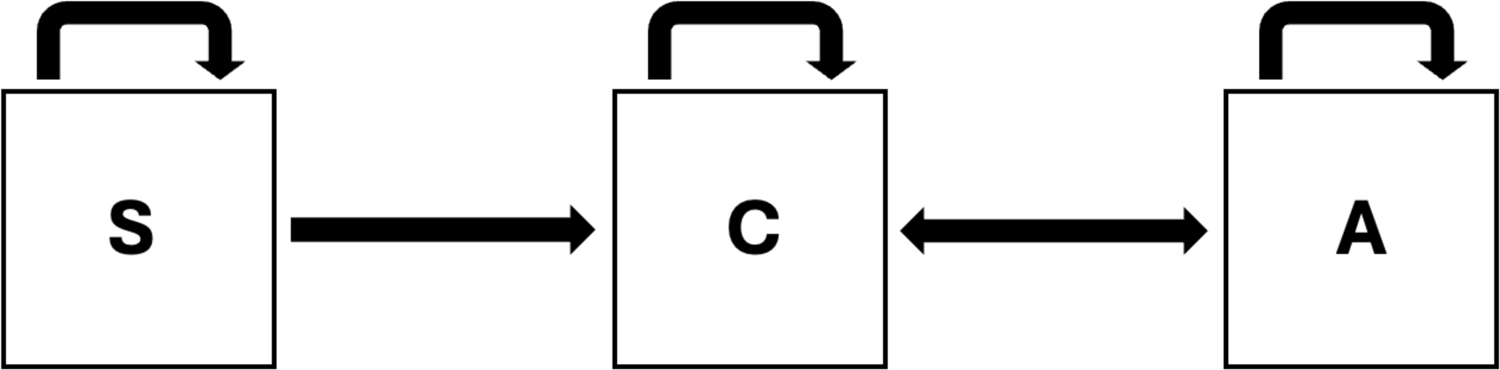
State transition model

Individual state transitions were computed separately for each 6-year molar, selected as these teeth are often the focus of school-based caries prevention. Following tooth-level analyses, we computed person-level transitions as an aggregate of all 6-year molars and first and second bicuspids over a period of one and two years. For this whole-mouth transition, any teeth with caries would result in a transition from either sound to carious or arrested to carious. If all previously carious teeth were arrested at any follow-up observation, a fully arrested state would be achieved. For instances where teeth were missing, the state from the previous observation was maintained. Analysis was conducted using R v3.3.0.

## Results

The total number of enrolled participants in the CariedAway trial was 7418 (Figure 2). After removing those who did not receive silver diamine fluoride (3679) and did not complete at least two follow-up observations (2387), the analytic sample consisted of 1352 children. The baseline prevalence of decay for the analytic set was 28.5%, and the baseline sealant prevalence (on any tooth) was 8%. Approximately 47% of the sample was male, predominately from Hispanic/Latino (59%) or black (21%) race/ethnicity. The average age at baseline was 6.44 years (SD=1.3).

**Figure 2:**
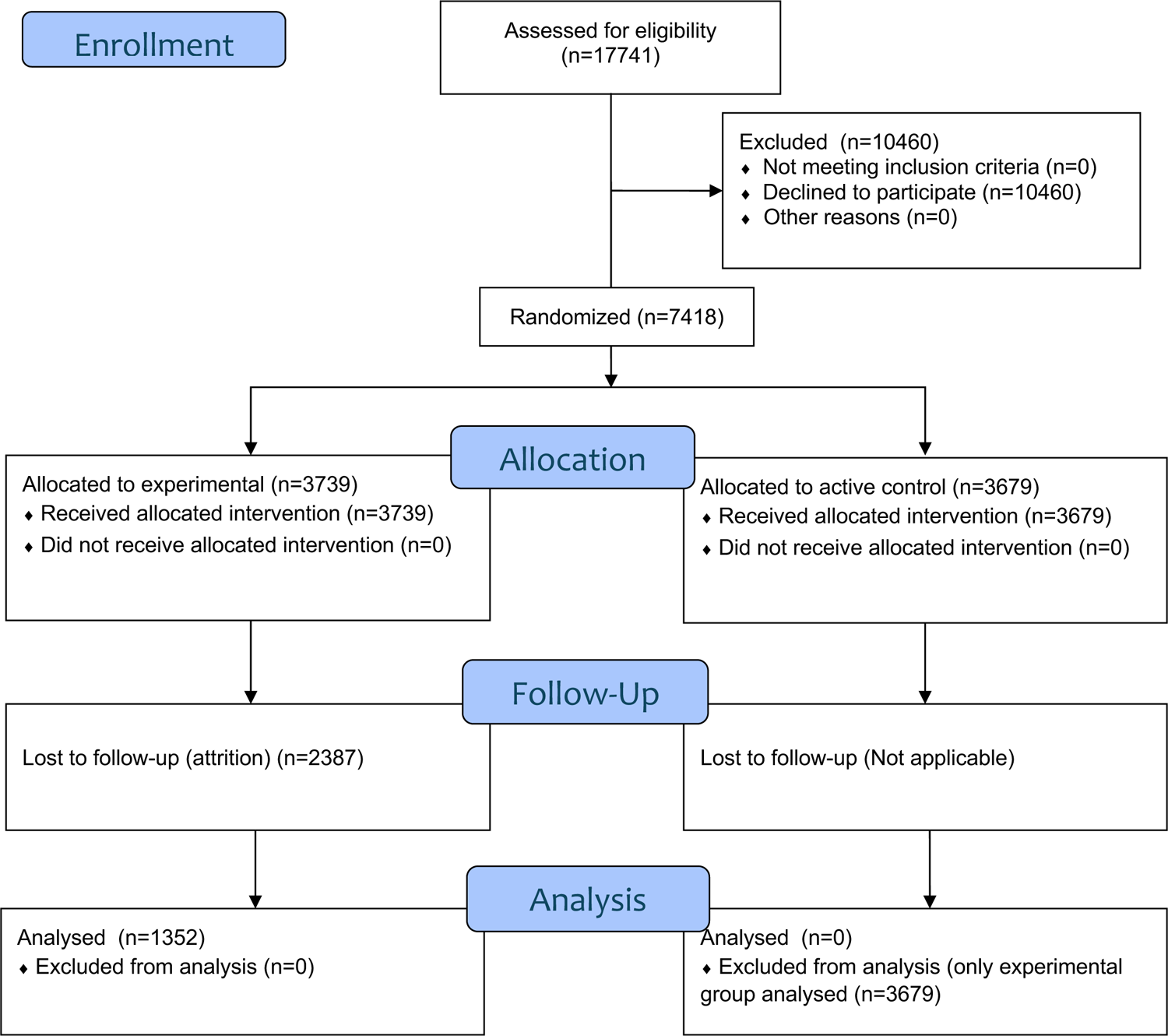
Study enrollment flowchart

For state transitions of 6-year molars (Table 2), probabilities for sound to carious transitions were 0.0037 (95% CI = 0.001, 0.006), 0.0022 (95% CI = 0.001, 0.004), 0.0058 (95% CI = 0.003, 0.009), and 0.007 (95% CI = 0.005, 0.011), while the initial probability of transitioning from carious to arrested ranged from 0.269 (95% CI = 0.125, 0.400) to 0.382 (95% CI = 0.255, 0.510). Once arrested, the probability of remaining in the arrested state (secondary prevention) ranged from 0.44 (95% CI = 0.287, 0.993) to 0.88 (95% CI = 0.633, 0.997).

**Table 1.** CariedAway study enrollment.

| Table 1 – CariedAway study enrollment |  |  |  |  |  |  |  |  |
| --- | --- | --- | --- | --- | --- | --- | --- | --- |
|  | Overall |  | SDF Group<br>(Enrolled) |  | SDF<br>(Analyzed) |  | Sealant/ART<br>Group |  |
|  | N | % | N | % | N | % | N | % |
| Participants | 7418 | 100 | 3739 | 50.4 | 1352 | 100 | 3679 | 49.6 |
| Baseline decay | 1980 | 26.69 | 1016 | 27.17 | 385 | 28.48 | 964 | 26.2 |
| Sex (male) | 3412 | 46 | 1785 | 47.74 | 637 | 47.12 | 1627 | 44.24 |
| Race/Ethnicity |  |  |  |  |  |  |  |  |
| Hispanic/Latino | 3648 | 49.18 | 1766 | 47.23 | 793 | 59.05 | 1882 | 51.16 |
| Black | 1246 | 16.8 | 650 | 17.38 | 286 | 21.30 | 596 | 16.2 |
| White | 153 | 2.06 | 86 | 2.3 | 35 | 2.61 | 67 | 1.82 |
| Asian | 125 | 1.69 | 88 | 2.35 | 36 | 2.68 | 37 | 1.01 |
| More than one | 114 | 1.54 | 67 | 1.79 | 21 | 1.56 | 47 | 1.28 |
| Other | 90 | 1.21 | 56 | 1.5 | 33 | 2.46 | 34 | 0.92 |
| Unreported | 2042 | 27.53 | 1026 | 27.44 | 139 | 10.35 | 1016 | 27.62 |
| Age at baseline | 7.58 | 1.90<br>(SD) | 7.55 | 1.92<br>(SD) | 6.44 | 1.32<br>(SD) | 7.63 | 1.88<br>(SD) |

**Table 2.** Tooth-level transition probabilities.

| Transition | Probability | 95% CI Lower | 95% CI Upper |
| --- | --- | --- | --- |
| 3<sub>11</sub> | 0.9960 | 0.9934 | 0.9985 |
| 3<sub>12</sub> | 0.0037 | 0.0014 | 0.0058 |
| 3<sub>13</sub> | 0.0007 | 0.0001 | 0.0012 |
| 3<sub>22</sub> | 0.7306 | 0.5996 | 0.8754 |
| 3<sub>23</sub> | 0.2694 | 0.1246 | 0.4004 |
| 3<sub>32</sub> | 0.1978 | 0.0063 | 0.4899 |
| 3<sub>33</sub> | 0.8022 | 0.5101 | 0.9937 |
| 14<sub>11</sub> | 0.9974 | 0.9948 | 0.9991 |
| 14<sub>12</sub> | 0.0022 | 0.0007 | 0.0043 |
| 14<sub>13</sub> | 0.0005 | 0.0002 | 0.0009 |
| 14<sub>22</sub> | 0.6806 | 0.5338 | 0.8140 |
| 14<sub>23</sub> | 0.3194 | 0.1860 | 0.4662 |
| 14<sub>32</sub> | 0.1205 | 0.0029 | 0.3672 |
| 14<sub>33</sub> | 0.8795 | 0.6328 | 0.9971 |
| 19<sub>11</sub> | 0.9926 | 0.9885 | 0.9957 |
| 19<sub>12</sub> | 0.0058 | 0.0034 | 0.0091 |
| 19<sub>13</sub> | 0.0017 | 0.0009 | 0.0025 |
| 19<sub>22</sub> | 0.6178 | 0.4902 | 0.7454 |
| 19<sub>23</sub> | 0.3821 | 0.2546 | 0.5098 |
| 19<sub>32</sub> | 0.2604 | 0.0056 | 0.5436 |
| 19<sub>33</sub> | 0.7396 | 0.4564 | 0.9944 |
| 30<sub>11</sub> | 0.9905 | 0.9867 | 0.9939 |
| 30<sub>12</sub> | 0.0074 | 0.0048 | 0.0113 |
| 30<sub>13</sub> | 0.0020 | 0.0012 | 0.0029 |
| 30<sub>22</sub> | 0.6781 | 0.5629 | 0.7921 |
| 30<sub>23</sub> | 0.3219 | 0.2079 | 0.4371 |
| 30<sub>32</sub> | 0.5629 | 0.0073 | 0.7132 |
| 30<sub>33</sub> | 0.4371 | 0.2868 | 0.9927 |

At the subject level (Table 3), the probability of remaining in a sound state (primary prevention) after one and two years was 0.916 (95% CI = 0.905, 0.925) and 0.840 (95% CI = 0.818, 0.855), respectively, for an annual prevention rate of approximately 7% and 12%. The probability of remaining in a carious state (any 6-year molar, 1^st^ bicuspid, or 2^nd^ bicuspid with caries, regardless of the states of the others) was 0.70 and 0.58 after one and two years. Once in a completely arrested state (no caries on any included teeth), the probability of reverting back to a carious state was 0.276 (95% CI = 0.222, 0.324) and 0.395 (95% CI = 0.530, 0.680) after one and two years, respectively. Predicted versus observed probability plots for each subject-level transition are shown in Figure 3, with tooth-level plots available as supplementary material.

**Figure 3:**
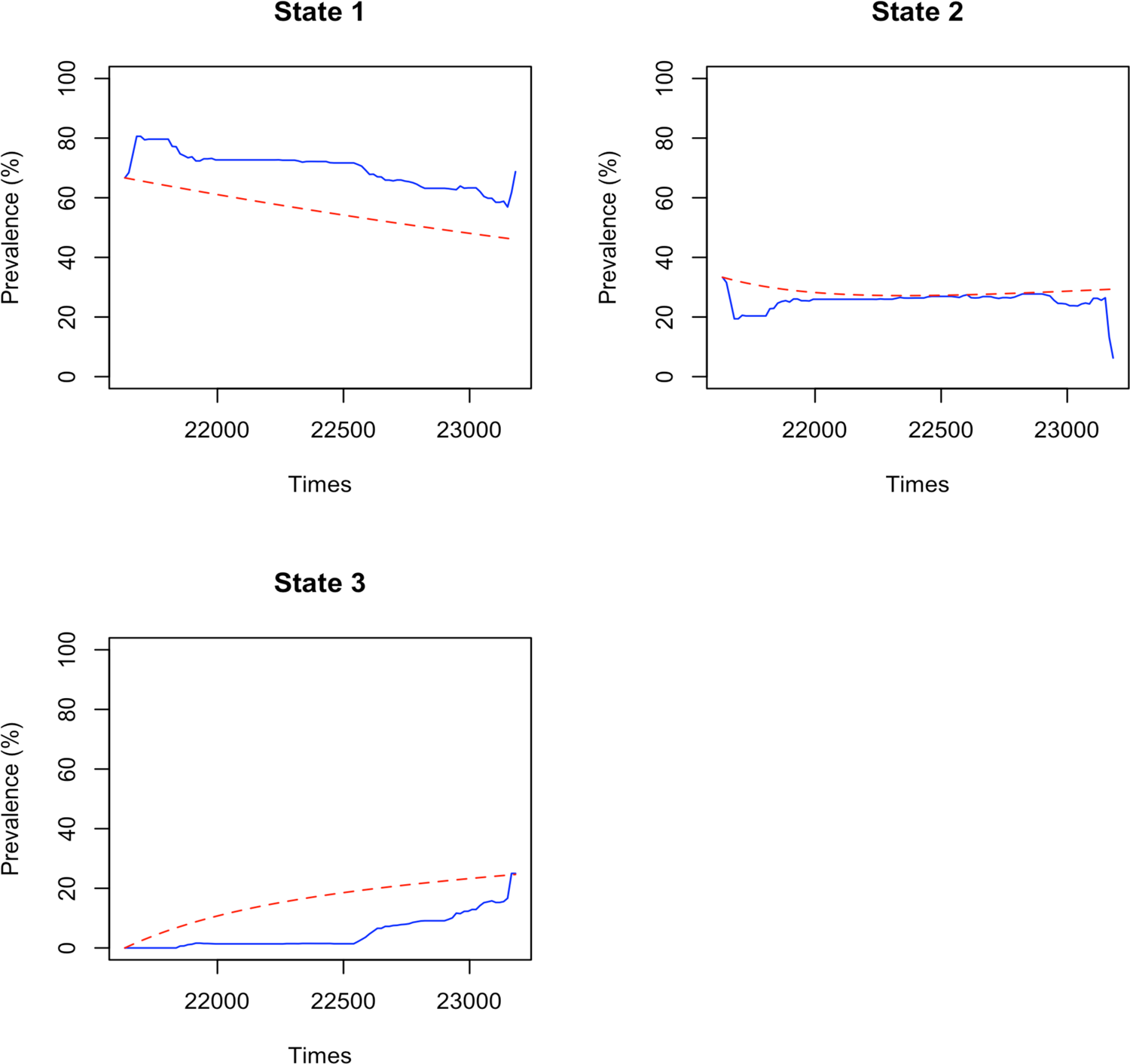
Predicted versus observed probabilities for person-level transition

**Table 3.** Person-level transition probabilities.

| Transition | Probability | 95% CI Lower | 95% CI Upper |
| --- | --- | --- | --- |
| *1-year* |  |  |  |
| M<sub>11</sub> | 0.9164 | 0.9054 | 0.9250 |
| M<sub>12</sub> | 0.0696 | 0.0619 | 0.0787 |
| M<sub>13</sub> | 0.0141 | 0.0118 | 0.0162 |
| M<sub>22</sub> | 0.7077 | 0.6783 | 0.7360 |
| M<sub>23</sub> | 0.2923 | 0.2640 | 0.3217 |
| M<sub>32</sub> | 0.2762 | 0.2215 | 0.3242 |
| M<sub>33</sub> | 0.7238 | 0.6759 | 0.7788 |
| *2-year* |  |  |  |
| M<sub>11</sub> | 0.8397 | 0.8179 | 0.8553 |
| M<sub>12</sub> | 0.1168 | 0.1045 | 0.1345 |
| M<sub>13</sub> | 0.0434 | 0.0379 | 0.0492 |
| M<sub>22</sub> | 0.5816 | 0.5446 | 0.6202 |
| M<sub>23</sub> | 0.4184 | 0.3798 | 0.4554 |
| M<sub>32</sub> | 0.3954 | 0.3203 | 0.4698 |
| M<sub>33</sub> | 0.6046 | 0.5302 | 0.6797 |

*M* = whole mouth transition, reflecting any change in 6-year molars, 1<sup>st</sup> bicuspid, and 2<sup>nd</sup> bicuspid

## Discussion

In this analysis of high-risk, low access children enrolled in a school-based clinical trial of caries prevention using silver diamine fluoride and fluoride varnish, the person-level transition probabilities from healthy to carious states were 0.07 and 0.12 for one and two-year periods, respectively. Once individuals with caries were in a fully arrested state, there was a 72% chance of remaining arrested after one year and a 60% chance after two years, suggesting high success rates of both preventing and managing dental caries.

All first permanent molars had high probabilities of remaining sound following initial treatment (> 0.99), while the first permanent molar in the lower left quadrant had the highest rate of arrest failure (0.437). This may be due to a higher overall risk of caries in lower molars, which is similar to data reported from epidemiologic studies ^19^ and clinical trials of SDF ^20^. Additionally, prior analysis of longitudinal caries risk estimated probabilities for transitioning from caries-free to caries-active ranging from 0.0046 to 0.047 ^21^, and treatment with fluoride varnish resulted in transition probabilities of 2.5% from sound to enamel caries and 0.3% from sound to dentine caries ^22^. Our results indicate similar transitions for high-risk children receiving school-based caries prevention.

As many chronic diseases follow a staged process, use of multi-state Markov models can effectively describe the course of a disease ^23^. The present application also highlights the natural progression of disease incidence and management in school-based caries prevention, which results in intermittently spaced observations of a continuous-time caries process. In these situations, providers are likely to observe mixtures of sound, carious, and arrested dentition when using an intervention with both preventive and control properties, such as silver diamine fluoride or dental sealants with atraumatic restorations, both of which have been used in school-based caries prevention ^9,14^. Additionally, other research has used semi-parametric multi-state frailty models in the analysis of clustered survival data with interval censoring, which may have multiple benefits for school caries prevention ^24^. Regardless of approach, modeling disease transitions at both the tooth and child levels can be useful for targeted planning of prevention in schools and minimize educational disruptions.

This study benefitted from using data from a randomized trial of school-based caries prevention that included a diverse mix of participants, with particular focus on those traditionally underserved in oral health. However, there are notable limitations. Dental caries severity was based off of clinical diagnosis following an ICDAS score of 5 or 6. While this supports a distinct state transition, it ignores potentially interesting earlier stages such as localized enamel breakdown due to caries. As this was also a diagnostic criterion that resulted in a recorded state of either carious or not carious, we were further unable to differentiate between adjacent stages of transitioning from ICDAS 5 to ICDAS 6, which would inform a more granular understanding of the caries process in school-based prevention. Furthermore, for analysis of 6-year molars, models could not account for possible intraoral correlation which may result in the estimated probabilities being attenuated. We attempted to address this through a person-level analysis, which is useful for planning and implementation of school caries prevention as the primary cost and time considerations for care is predominantly due logistics (e.g., estimating needed supplies, visiting a school, setting up mobile dental chairs, and identifying children in need), and not on the treatment itself. However in this approach, state transitions from arrested to carious no longer reflect only secondary prevention, as new carious teeth would force the participant into a carious state when it may have been due to a sound to carious (from another tooth) transition. Despite this limitation, this approach can still be useful for program planning, as disease prevalence is the fundamental concern rather than the underlying process.

In conclusion, use of silver diamine fluoride for school-based caries prevention demonstrated high probabilities of primary prevention and caries management over one to two-years. These results can better inform strategic implementation of a school SDF program and maximize resources to treat the most children in need.

## Data Availability

Data available: Yes
Data types: Data dictionary
How to access data: Data dictionaries will be available to interested researchers upon request to the authors (ryan.ru=@nyu.edu)
When available: beginning date: 06-01-2025
Supporting Documents
Document types: Informed consent form
How to access documents: Informed consent forms will be available to interested researchers upon request to the authors
When available: beginning date: 06-01-2025
Additional Information
Who can access the data: Interested researchers upon request to the authors
Types of analyses: For any purpose
Mechanisms of data availability: After approval of a proposal and a signed data access agreement.

## Supplementary Material

**Supplementary Figure 1:**
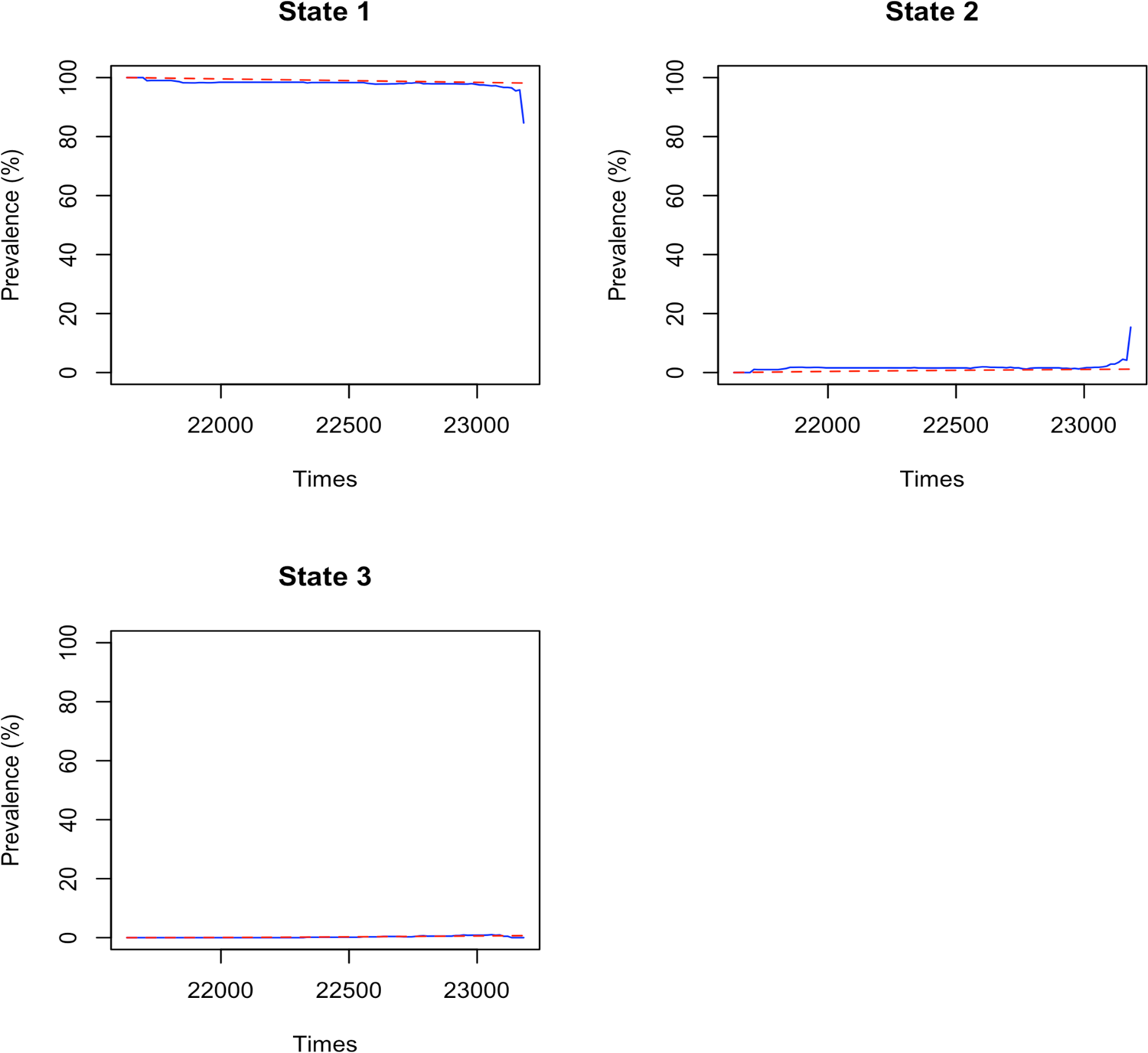
Predicted versus observed probabilities for tooth 3

**Supplementary Figure 2:**
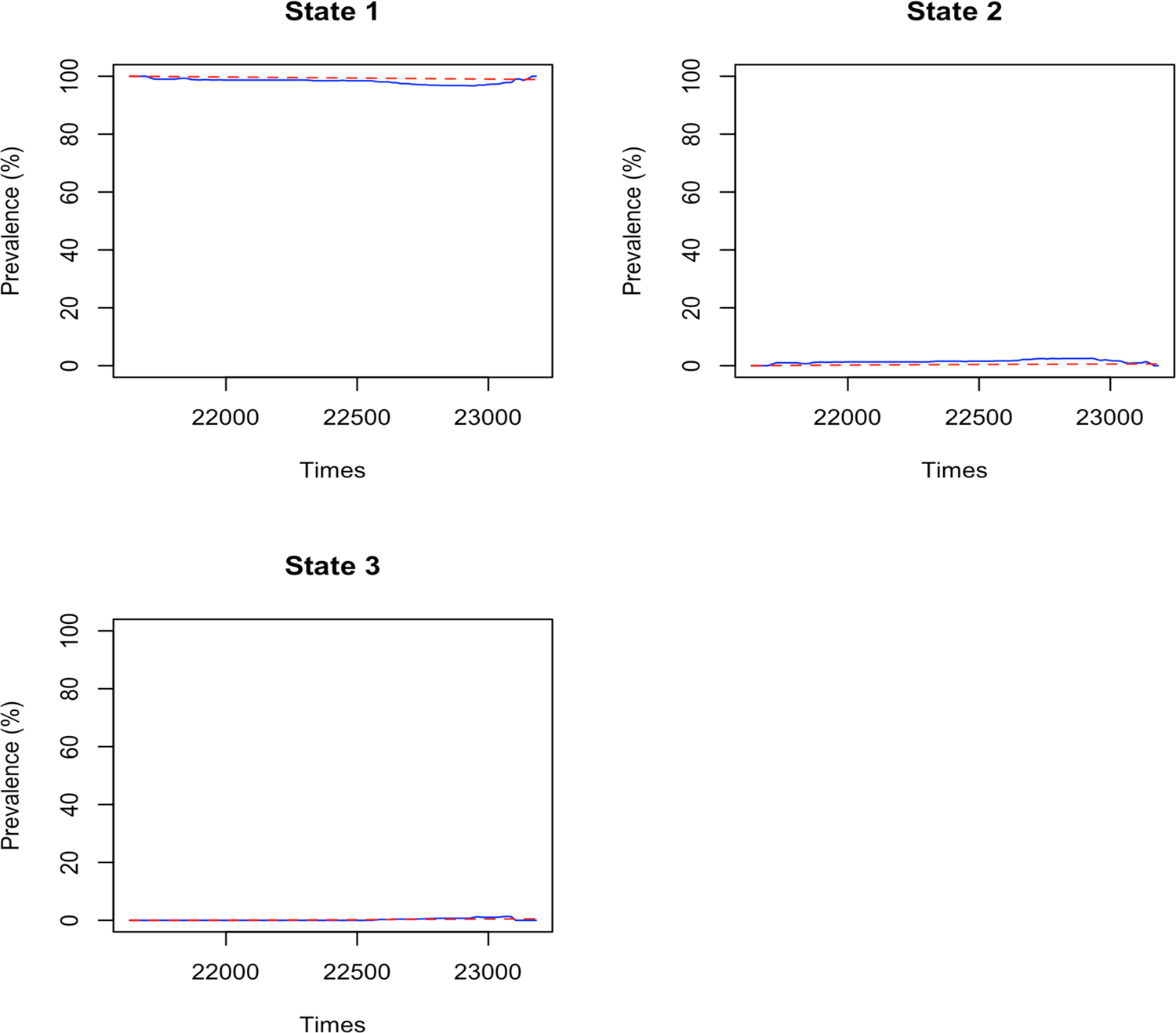
Predicted versus observed probabilities for tooth 14

**Supplementary Figure 3:**
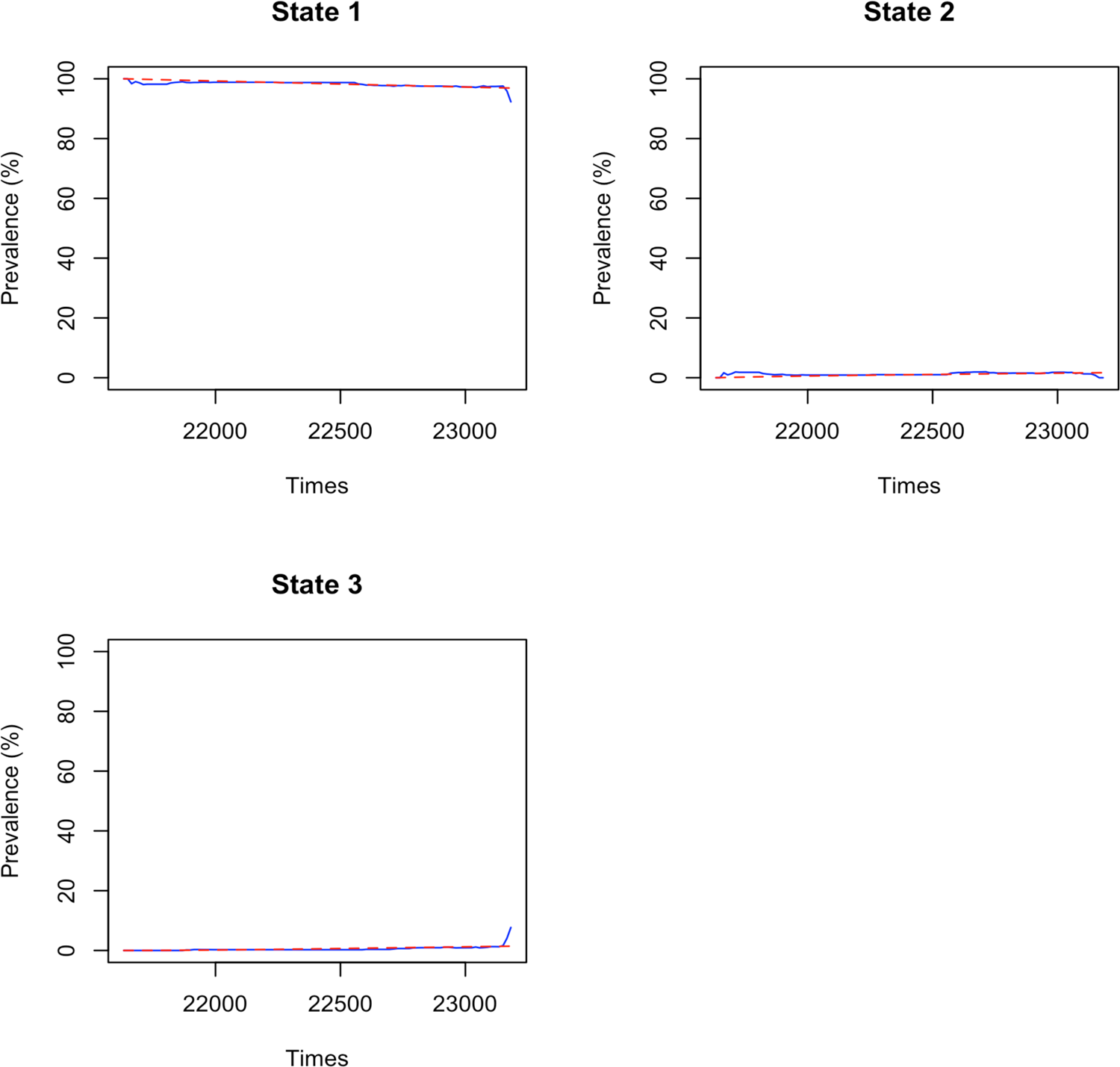
Predicted versus observed probabilities for tooth 19

**Supplementary Figure 4:**
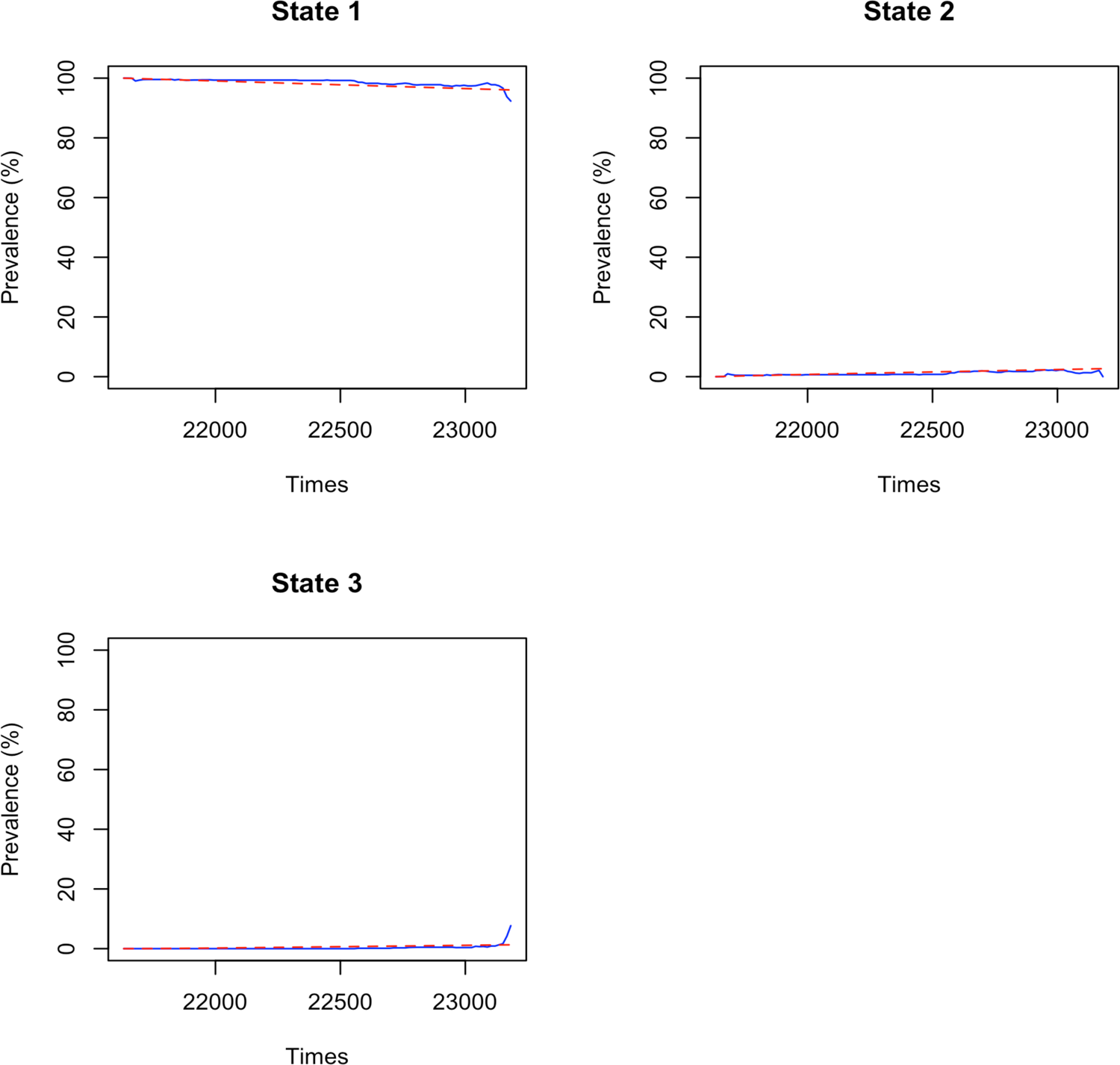
Predicted versus observed probabilities for tooth 30

## Notes

### Competing Interest Statement

The authors have declared no competing interest.

### Clinical Trial

NCT03442309

### Funding Statement

Research presented in this report was funded by the Patient-Centered Outcomes Research Institute (PCS-160936724).

### Author Declarations

The CariedAway study received approval from the New York University School of Medicine Institutional Review Board (#i7-00578).

